# Knowledge Mapping of Nigeria’s Scientific Contribution to Antimicrobial Resistance Research: A visualized investigation using VOS viewer and Cite Space

**DOI:** 10.1101/2022.09.20.22280150

**Authors:** Shuaibu Suleiman Adeiza, Gazali Shuaibu Mohammed, Bello Abdulmalik Shuaibu

## Abstract

**Objective:** A comprehensive overview of the research output, including conceptual and temporal trends, is required to address the growing AMR burden in Nigeria, which was previously lacking. This study uses bibliometric analysis to show this.

**Methods:** Data from a query of AMR articles in the Dimensions database (PubMed included) were subjected to bibliometric analysis (1972–2022). Bibliometric indicators, topic networks were established and topic trends were analysed by frequency, relevance scores, and trends over time.

**Results:** A total of 0.2% of the papers on AMR published worldwide were written by authors or institutions from Nigeria. In 2021, publications grew by 13.6%, highlighting the field’s increased vigour. The vast majority of publications (57.18%) were in the field of Medical and Health Sciences, with 2428 papers. The top journal for AMR papers with at least one Nigerian institution affiliation was African Journal of Clinical and Experimental Microbiology, which published 1.8% of these papers. Antimicrobial stewardship, Clinical and laboratory practices on AMR, Public health implications, Traditional and molecular methods, and Phytomedicine and drug discovery were prominent areas of focus for Nigerian researchers.

**Conclusion:** This study provides a comprehensive overview of the AMR research output, highlighting the need of increased research capacity to address the burden of AMR in Nigeria. Because Nigeria is overly reliant on conventional methods of discovery and development, innovation is essential for the future of antibiotics.

## Introduction

Microbes can become resistant to antimicrobials through mutation or adaptation, which renders treatments ineffective. This phenomenon is known as antimicrobial resistance (AMR)[1]. The World Health Organization (WHO) estimates that Antimicrobial Resistance kills over 700,000 people across the globe yearly, an estimate that may well soar to 10 million by the year 2050[2]. According to a needs assessment on antimicrobial use and resistance in Nigeria published in 2017 by the federal ministries of health, agriculture, and the environment, certain antimicrobial agents have increased resistance rates of 70% to 100% in certain states[3]. In a country where infectious diseases are a major cause of morbidity and mortality, Nigeria considers AMR a priority on the national public health agenda[3]. The Nigerian response to antimicrobial resistance led by the Nigeria Centre for Disease Control (NCDC) began in 2016 following a situational analysis that investigated common antimicrobial-resistant pathogens recovered from hospitals, animal sources, agricultural and environmental sources[4]. Overall, the National Action Plan (NAP) strives to address five key pillars in accordance with the WHO Global Action Plan on Antimicrobial Resistance[5]. The fundamentals entail boosting knowledge and awareness of AMR among health personnel and the general public; developing a One Health surveillance system; strengthening prevention efforts, control, and biosecurity; promoting rational use of antimicrobials and access to quality medications; and conducting research into antimicrobial alternatives, new diagnostics, and drug discovery[4]. The problem of AMR in Nigeria is multisectoral. The ease with which antibiotics are available and can be purchased without the need for a prescription thrives in an environment where the antibiotic market is poorly regulated. The problem of improper disposal of expired antibiotics and left-over medicines by the general public, which mostly end up in conventional waste bins and dumps due to lack of awareness on the part of the users and weak enforcement on the part from regulatory authorities[6]. The problem of indiscriminate use impacts human health and animal welfare alike [7]. In the human health sector, many incidences of fever are addressed empirically with antibiotics[8]. Although a few antimicrobials are prescribed by a doctor, patients and their families frequently press healthcare professionals for antimicrobials and, in most cases, self-medicate with antimicrobials even for mild viral infections[9]. Even with the Nigerian Veterinary Council’s warnings against excessive use of antimicrobials in the animal health sector, several farmers habitually add antimicrobials into animal feed or water to promote growth and, in the case of poultry farming, to boost egg size to ensure better profit. Several farmers in the country use antimicrobials in post-harvest storage to preserve the yield[10]. As a result, human and animal waste, particles from crop sprays, discharge from health facilities, pharmaceutical companies, and farmlands carrying antimicrobial-resistant pathogenic microbes and antimicrobial residues find their way into the ecosystem[11]. It thus results in successions of negative impact on food production, bodies of water, as well as fish farms[11]. Despite increased awareness of antimicrobial resistance, the general situation is deteriorating, and the scientific community and Institutions have endured in research and publications to fashion newer antimicrobial peptides (AMPs), antibiotic combinations, and monitoring systems to effectively control AMR[12]. The amount of reliance on data increases with the growth of scientific publications and information technologies in healthcare. These big data raise various issues to be resolved by innovative big data analytics, like Knowledge mapping via bibliometrics.

Knowledge mapping is an assessment tool that blends concepts and techniques from mathematics, visuals, and other disciplines with bibliometric methods[13]. Cite Space, and VOS viewer are the most popular bibliometric analysis tool for Knowledge mapping via bibliometrics[14]. Knowledge mapping has been applied in many research fields[13,15,16]. To date, a knowledge mapping analysis of AMR research contributions and inclinations in Nigeria has remained hidden. This study aimed at filling this gap. Understanding Nigeria’s contribution to AMR research requires quantifying and mapping AMR-related publications. This study examines the research conducted by institutions in Nigeria using the Dimensions database as a foundation. We focus on bibliometric elements like international collaboration, subject area classification, active authors and institutions, journals and highly cited papers. In this study, we generated visualized knowledge maps of AMR research in Nigeria and analysed scientific hotspots, emerging/future trends, prolific authors and key research institutions of momentous research activities in AMR, which we hope will provide clarity about the nation’s priority for AMR research; and serve as reference for future collaborative networks.

## METHODS

### Data acquisition and query criteria

Bibliographic data on antimicrobial resistance publications in Nigeria were retrieved from the Dimensions™ database on August 20^th^ 2022. The database is one of the most comprehensive for bibliometric and patentometric analysis. It contains millions of publications cited over 1.6 billion times linked with Patents,clinical trials,policy documents, datasets and grants [15,17]. This database is completely inclusive of PubMed, making it highly reliable. We used the search criteria: ‘antibiotics OR antifungal OR Antibacterial OR antimicrobial resistance AND Nigeria’ to spotlight research articles published between 1972 and 2022. The search was not limited by language, and it included all accessible data sets. Both title and abstracts were search to ensure subject-specific outcome for the benefit of accuracy. Publications indexed in the database that featured a minimum of one author connected to a Nigerian institution, including a university or organization, qualified for inclusion.

### Data processing and parsing

The dplyr and ggplot2 packages of R-studio v.4.1.2 (2022-02-16) was used to examine, parse and visualize bibliographic data. Imported raw files were cured of false positives and duplicates. Top 100 documents were arbitrarily selected per year and checked for presence of false positive results. A key term was ascribed to noun-phrases with similar meanings (e.g., “Antimicrobial sensitivity test “, and “AST” were merged to “Antimicrobial susceptibility test “). Multiple or similar keyword occurrences in a publication were consolidated as a single count. Network mapping and visualizations were rendered with Visualization of Similarities (VOS) Viewer software (Leiden University, Netherlands)[18].This method adopts diverse clusters and Color spectrum ; the bubble size symbolizes the total frequency, the colours indicate the cluster type, and the lines of conjunct (straight or curved) illustrates connectedness [19]. Indices like, scientific production, mean citations, authors productivity, affiliation and countries, preferred journals and keywords, were used to assess the present and likely future trends of AMR research in Nigeria [20,21]. The terms used in keyword clustering analysis were extracted from title and abstract fields of included bibliographic documents. copyright statements and Structured abstract labels were ignored in the analysis. Full counting method with minimum occurrence threshold of 20 (T≥20) was used. The top 60% of calculated relevance scores were used in the analysis.

In order to determine the burstness and centrality index of the bibliographic components, Cite Space (Version 6.1.R3 Basic) was used[22]. The following limits were set: time span (1972 to 2022), years per slice (one), pruning (pathfinder and Pruning Sliced Networks), selection criteria (g-index), link strength (cosine) and scope (within slices) and other default settings.

Furthermore, the top cited articles were also extracted and sorted by field citation ratio. The Field Citation Ratio (FCR) and Relative Citation Ratio (RCR) are citation-based measures of an articles’ scholarly impact[23]. FCR was determined by dividing the number of citations received by an article by average number of citations received by articles published within the same year and Fields of Research (FoR) class. While the RCR is measured as the number of citations received by a paper adjusted to the citation count garnered by National Institute of Health (NIH) funded publications in the same research field and year[23].

## RESULTS

### Nigeria’s annual contribution in AMR Research

Nigerian authors and institutions have played a key role in production of a total of 4,246 papers on antimicrobial resistance. As noted in Figure 1, AMR-related research in Nigeria have shown an annual upward tendency that peaked in 2021(13.6%). Based on our research, there are 2,176,583 publications on AMR globally and Nigeria have contributed at least 0.2% of that number. Table S1 shows the distribution of scientific output by field of research; Medical and Health Sciences (n = 2428, 57.18%), Biological Sciences (n = 806, 18.98%), Agricultural and Veterinary Sciences (n =409, 9.63%).

### Influential Contributor Analyses

#### 1. Authors and Co-Cited Authors contributing to AMR research

A total of 14391 authors were involved in AMR research. OKEKE IN published the most papers indexed in Dimension database (n = 31), followed by LAMIKANRA A (n = 28), and OLAYINKA BO (n = 27) (Figure 1). The top ranked author (cited) by centrality is BAUER AW (1985) with centrality of 0.13 followed by Okeke IN (2000), with centrality of 0.07 (Figure 3). The co-authorship network map was constructed authors (n = 1000) who had published at least five papers on AMR (Figure 4). Authors that share the same colour space represent the same cluster. There were active collaborations in AMR research in Nigeria, especially among authors in the same cluster, such as OKEKE IN AND LAMIKANRA A (green), SOWUNMI A and ODUOLA AMJ (coffee brown), ILIYASU G and HABIB AG (lime green), OLAYINKA BO AND ONAOLAPO JA (yellow) etc.

**Figure 2:**
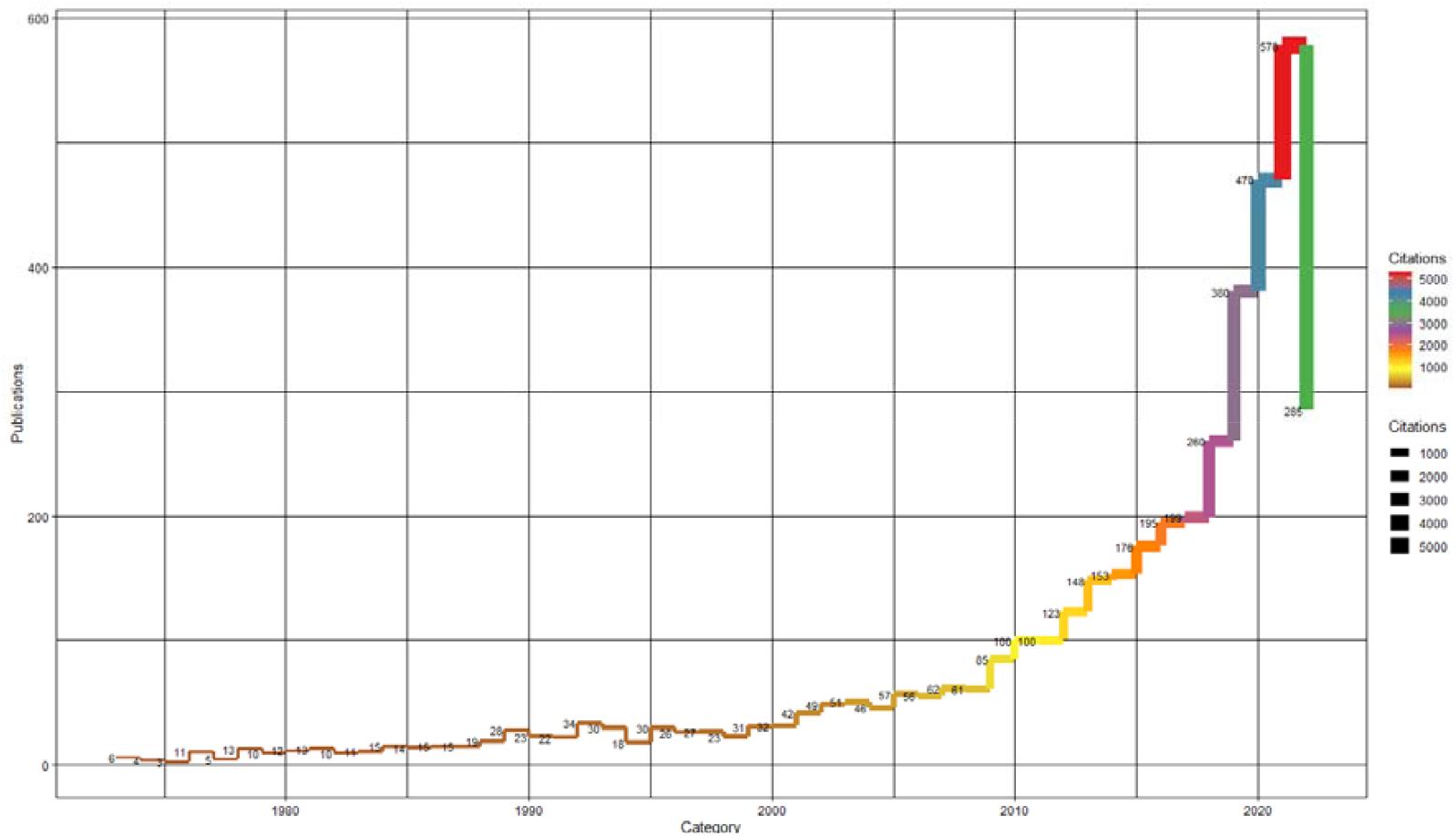
Nigeria’s annual contribution to AMR publication growth.

**Figure 3:**
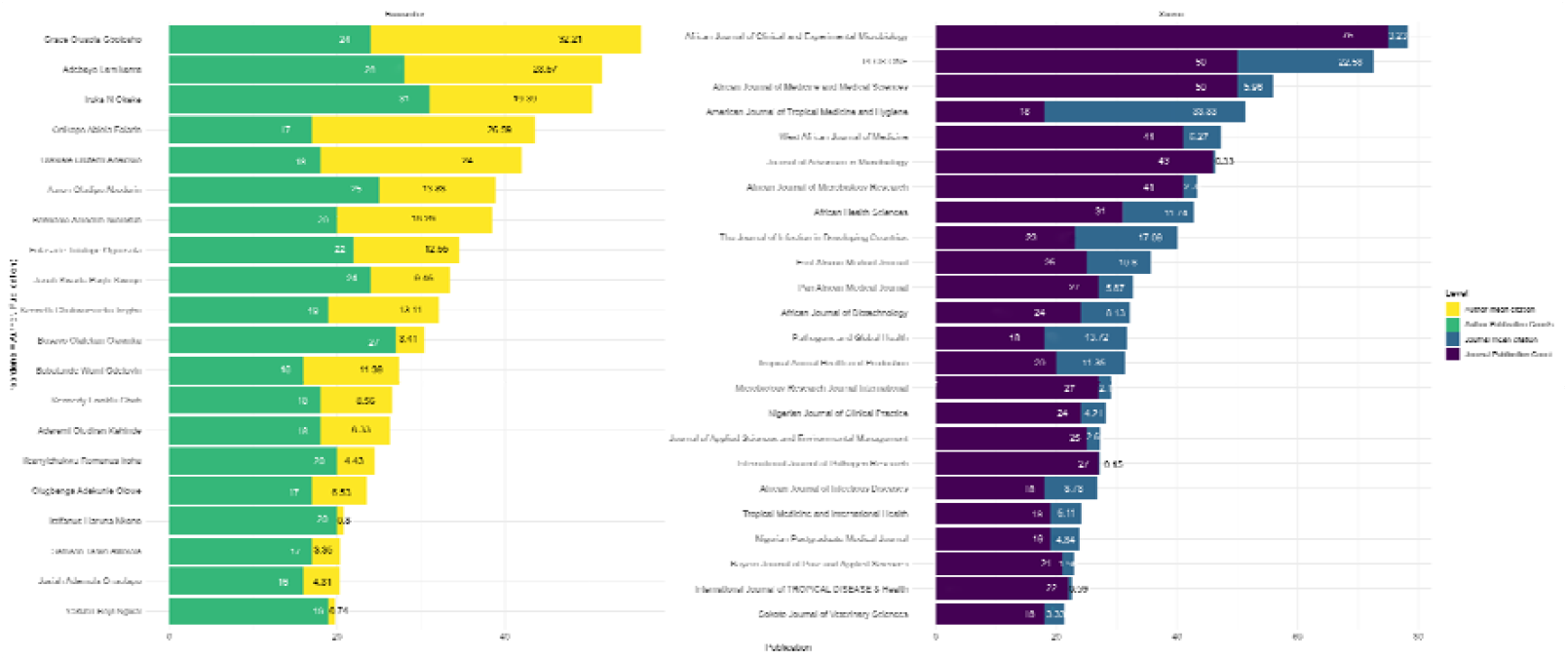
The top Authors and Journals of AMR research in Nigeria

**Figure 3:**
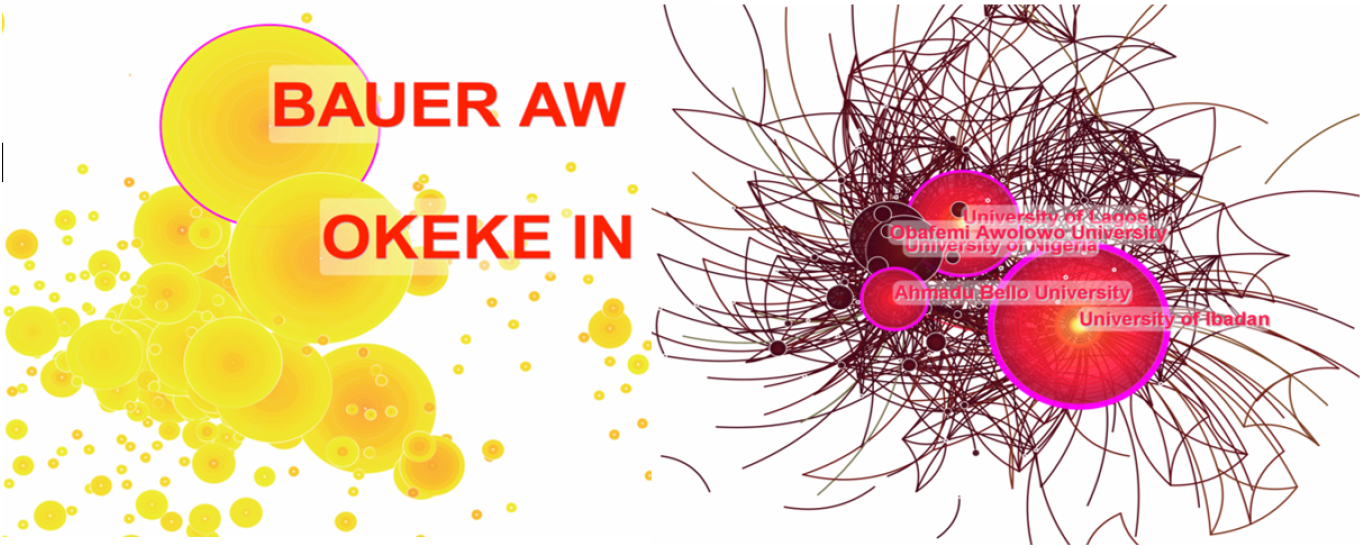
Centrality of top Universities (Right) and Authors (Left). Notes: each node in the network represents cited Author /University, the number of papers published by the author is represented by the size of the nodes, and the stroked rings represent the centrality index.

**Figure 4:**
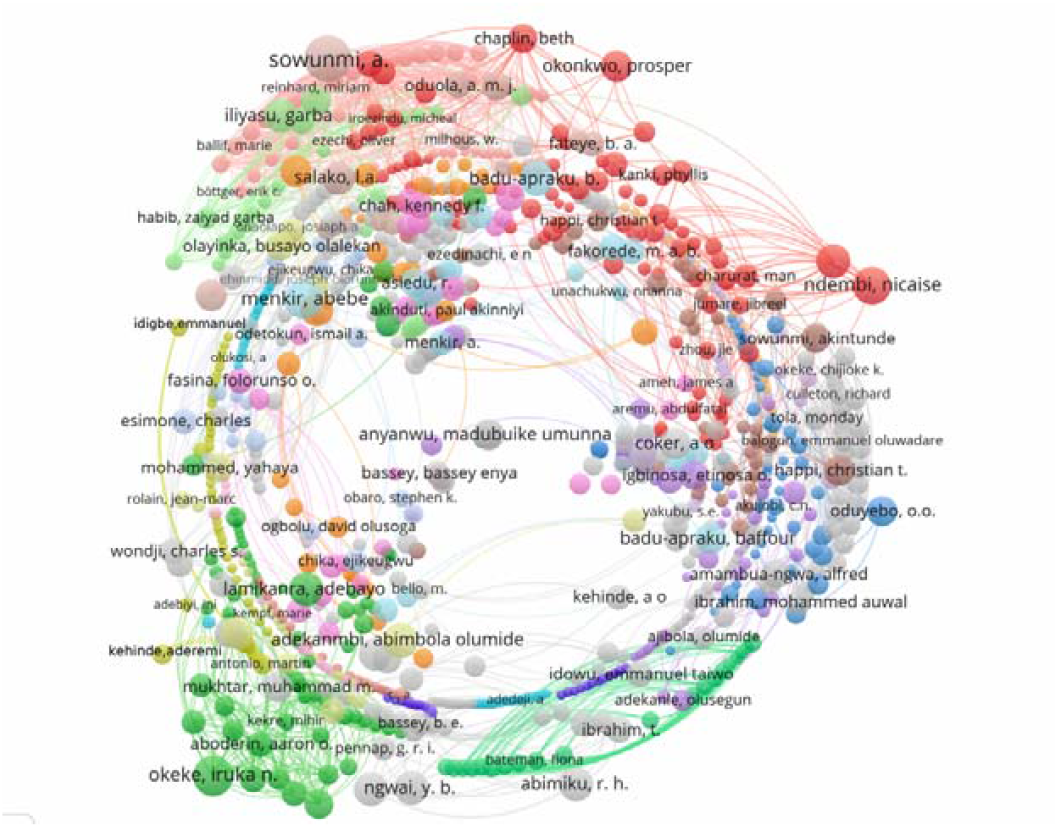
The co-authorship map of 1000 authors of authors in AMR research domain (T≥5). Notes: The size of node reflects the author’s co-occurrence frequencies, the link specify the co-occurrence association between authors, and the same colour of node represent the same cluster. In the figure there are 74 clusters, 7063 links and a total link strength of 10240.

Authors with citation bursts are defined as those that are cited frequently over a while. Through detecting bursts, Table 1 lists the top Authors of publications on AMR with the strongest citation bursts. COKER A on the top of the list with the maximum burst duration of 15 years and strength of 4.54, followed by OGUNSOLA F with a 14-year citation burst with 4.45 burst strength. NGWAI Y had a minimum burst duration of 1 year and the highest strength of 8.7. Recently published authors of AMR research with the strongest burst were NGWAI Y (8.7), OLAYINKA B (8.53) and OKEKE IN (7.27). This shows that their work may have formed a hot and leading topic. The table also show a list the top cited authors of publications in AMR with the strongest citation bursts. AKO-NAI AK on the top of the list with the maximum burst duration of 18 years and strength of 6.5, followed by TENOVER FC A with an 8-year citation burst with 5.86 burst strength. OKEKE IN had the strongest burst 11.6 among cited authors.

**Table 1:**
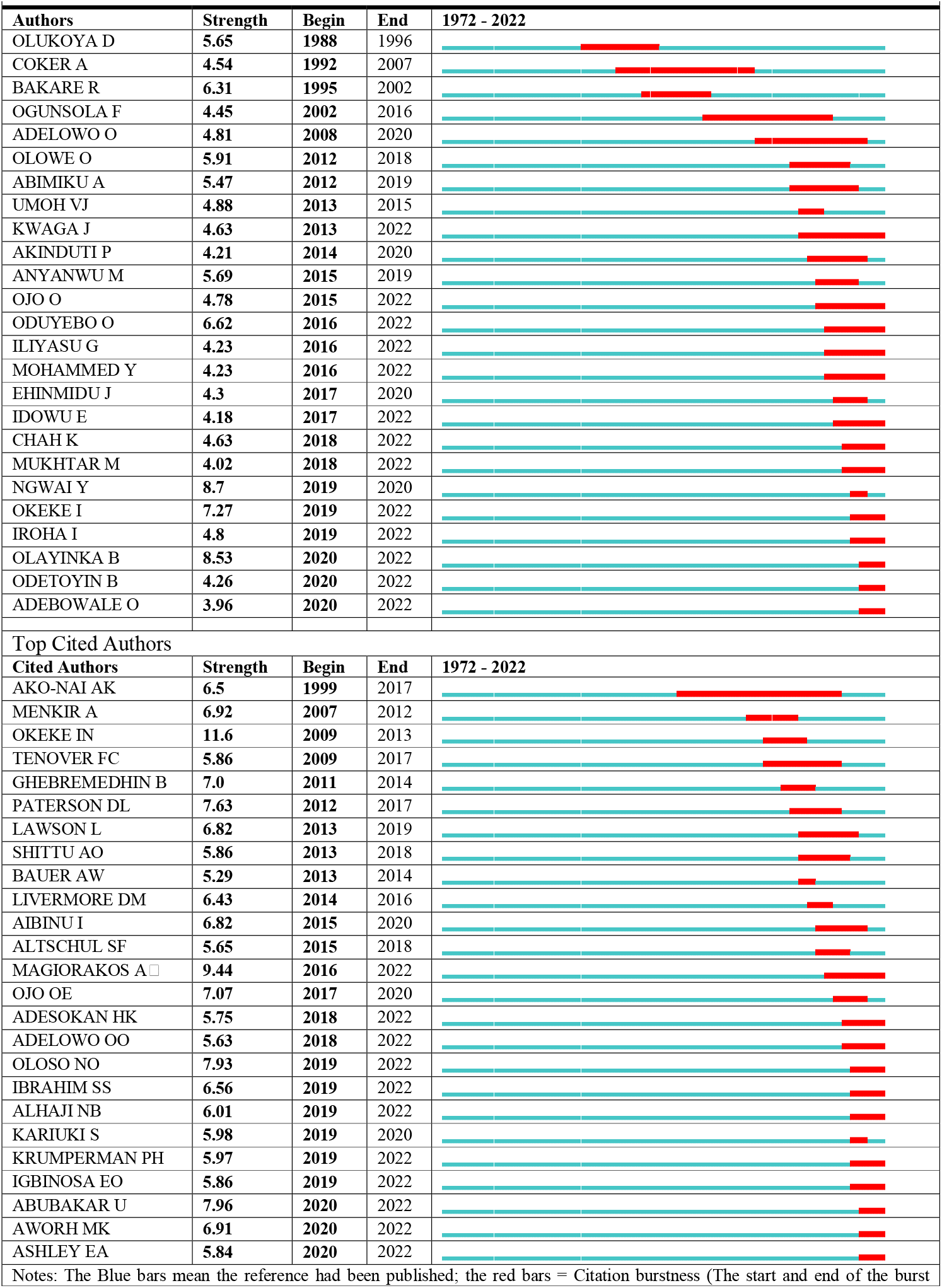
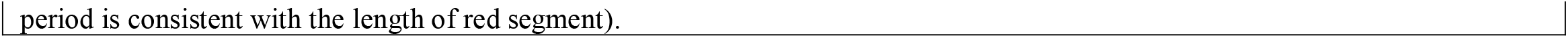
Top25 Authors and Cited Authors in AMR research with the Strongest Citation Bursts (Sorted by Beginning year)

#### 2. Contributions and Cooperation Network Among Institutions/Organizations

The top ranked institutions/organizations in AMR research collaboration by centrality are University of Ibadan (1972), with centrality of 0.33. The second one is Ahmadu Bello University (1972), with centrality of 0.14. The third is University of Lagos (1985), with centrality of 0.13. The 4th is International Institute of Tropical Agriculture (1975), with centrality of 0.13. The 5th is Obafemi Awolowo University (1982), with centrality of 0.10 (Figure 3). Figure 5, depicts the cooperation network among Nigerian institutions/organizations on AMR research, where the nodes represent the institutions/organizations, and the sizes denote number of publications. The lines between two institutions/organizations denote that they cooperate. The density of the links of an institutions/organization indicates the cooperation degree. The denser of the lines, the more collaboration for one institutions/organization with other institutions/organization. From the figure, University of Ibadan (n =366, links = 1586), University of Lagos (n =218, links = 1079), University of Nigeria (n =172, links = 430), Ahmadu Bello University (n =152, links = 647), Obafemi Awolowo University (n =141, links = 566) and Nigerian Institute of Medical Research (n =132, links = 883) have more lines. Therefore, they have more cooperation with other institutions/organizations.

**Figure 5:**
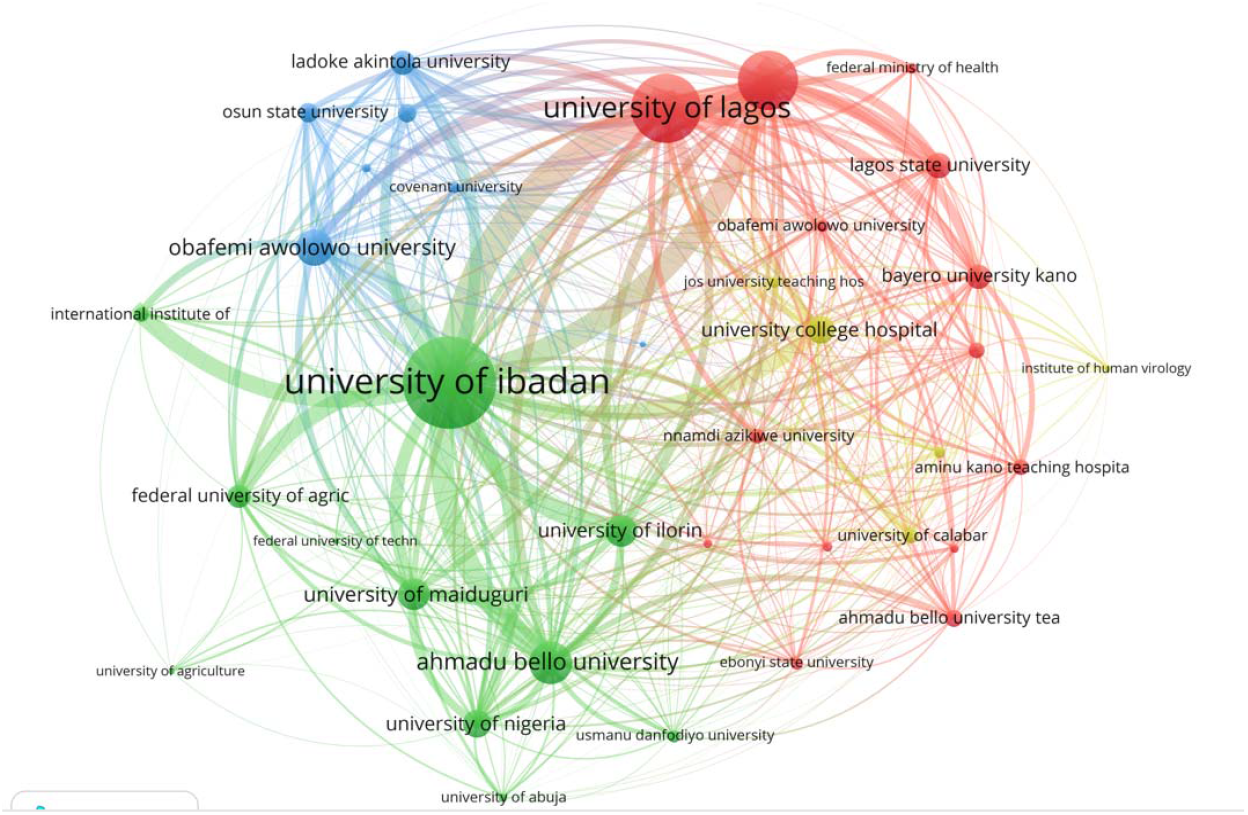
Institutional collaboration network: A minimum threshold of 20 (T = 20) documents per institution was set for inclusion. Notes: The size of the connecting lines, circles and colour vary based on total link strength and number of publications and cluster respectively. Normalization method used was Lin/log modularity and full counting method.

#### 3. Analysis of Top Journals publishing Nigeria’s contribution to AMR research

By analysing the distribution of journals accepting Nigeria’s contribution in this field, we could accurately identify the main part of the academic research, and papers published in such journals that could be supported by academic research. Figure 2 shows the top journals that published Nigeria’s contributions in AMR research. The number of publications in these journals was about 21.83% of the total number of contributions in this field. The number of articles published in African Journal of Clinical and Experimental Microbiology was the largest, with 75 articles, accounting for 1.8% of the total. Followed by PLoS One and African Journal of Medicine and Medical Sciences, with 50 articles each (1.2%). In terms of mean citation, American Journal of Tropical Medicine and Hygiene had the highest (33.3%).

#### 4. AMR Research themes and trends in Nigeria

Keywords are the condensation and reaction to the main content of the article, which can reflect the hot topic and the development trend related to the research field. We ran keyword co-occurrence network analysis with VOS viewer and arrived with a graph with 953 nodes and 6 Clusters (Figure 6). The keywords “*Escherichia coli*” and “*Staphylococcus aureus*” displayed the highest frequency of 881 and 540, respectively. Followed by the keywords “ESBL” (470), “*Salmonella*” (468), “tetracycline” (465) and “plasmid” (412). Thematic research group of focus by Nigeria authors are; Antimicrobial stewardship (25.50%), Clinical and laboratory (25.39%), public health implication (23.92%), traditional and molecular methods (16.79%) and phytomedicine and drug discovery (8.39%). Additional insights on the clusters are described below;

**Figure 6:**
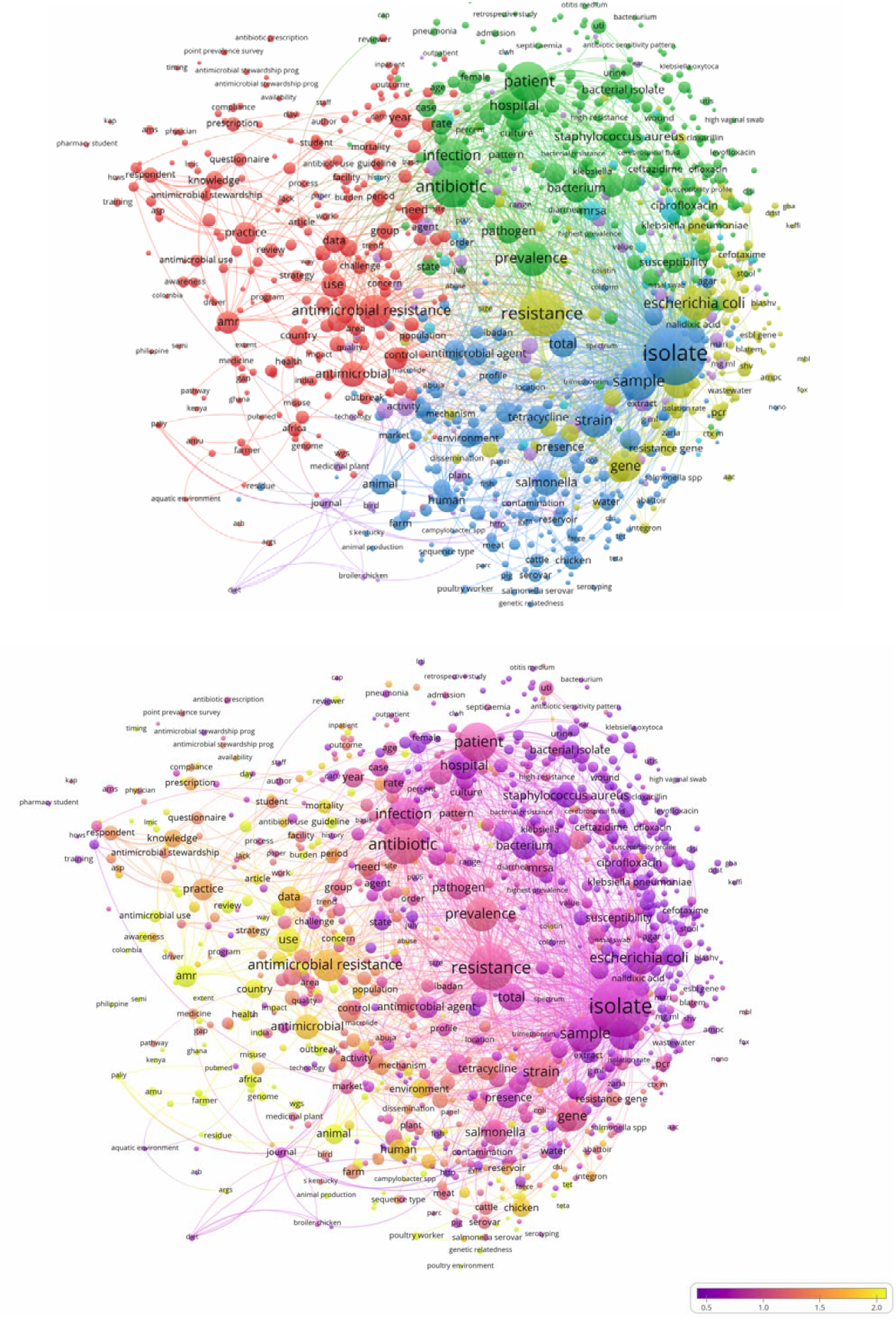
keyword co-occurrence network and clusters of AMR research (T≥40, contains 1481 items, 14 clusters and 449516 links; max lines = 200, total link strength = 5333437). Notes: The node and word size reflects the co-occurrence frequencies, the link indicate the co-occurrence relationship, and the same colour of node signify the same cluster. Network map of the trend topics according to the keywords used. Indicator shows the current publications from purple to yellow. More studies focused on vaccine, drug treatment and spike protein have been published recently. The size of the circles represents the frequency of appearance as the keywords. The distance between the two circles indicates their correlation.

#### a) Antimicrobial stewardship theme-based cluster (red, n= 243 items, 25.50%)

Antimicrobial stewardship, antibiotic prescription, antimicrobial resistance, aquatic environment, antimicrobial usage, attitude, awareness, burden, compliance, combination, evaluation, knowledge, middle income country, misuse, monitoring, mortality, observation, perception, point prevalence, practice, prescription, rational use, self-medication, standard, surveillance, survey, Systematic review, world health organization etc.

#### b) Clinical and laboratory theme-based cluster (green, n= 242 items, 25.39%)

Patients, outpatient, treatment, case, Acinetobacter baumanii, 30ug, Adult, age group, Aeromonas hydrophilia, amoxicillin, amoxiclav, ampicillin, antibiotic, antenatal clinic, antibiogram, antibiotics resistance profile, antibiotic susceptibility test, antibiotic resistance, Augmentin, azithromycin, bacillus, bacteriaemia, bacteria isolate, bacteria pathogen, bacterial infection, bacterial resistance, biofilm, blood agar, cefepime, cefixime, cephalosporin, cerebrospinal fluid, choice, cloxacillin, conventional biochemical test, cotrimoxazole, disk diffusion method, drug resistance, *Enterobacter* spp, first line drugs, gram stain, high resistance, high virginal swab, erythromycin, intensive care unit, *Klebsiella*, Kirby Bauer disc diffusion method, levofloxacin, microbiology laboratory, multi drug resistant, pattern, otitis medium, *Pseudomonas* spp, *Shigella* spp, standard microbiological techniques, samples, UTI, wound infection, swab etc.

#### c) Public health implication theme-based cluster (blue, n= 228 items, 23.92%)

Isolate, strain, gene, animal, human, Abattoir environment, antibiotics resistance gene, animal production, antimicrobial residue, aquaculture, biofilm formation, cattle, coliform, environmental sample, faecal sample, ESBL, farm, fluoroquinolones, food handlers, gastroenteritis, indiscriminate use, livestock, *Listeria spp*, meat sample, MLST, market, oxytetracycline, phenotypic characterization, poultry environment, pork, public health implication, raw meat, reservoir, resistance profile, *Salmonella sp*. salmonellosis, seafood, serotyping, whole genome sequencing, standard bacteriological method, *Vibrio* spp, significance, virulence gene, resistance genes etc.

#### d) Traditional and molecular methods cluster (yellow, n= 160 items, 16.79%)

Molecular characterization, resistance gene, molecular detection, multiple antibiotics resistance gene, amp gene, Betalactamase, *blaZ, ctxm, bla-shv, bla tem*, PCR, carbapenem, cefotaxime, CIP, clinical infection, clinical isolate, clinical sample, detection, plasmid profile, plasmid, *pmqr*, sequencing, dissemination, double disc synergy test, *E. coli*, ESBl, gram negative, integrons, laboratory standard institute, MDR, MARI, *mec*A, gene, pathogens of public health concern, methicillin resistant *Staphylococcus aureus*, nasal swab, vancomycin, carriage, etc.

#### e) Phytomedicine and drug discovery themed cluster (purple, n= 80 items, 8.39%)

Plant extract, aqueous extract, antibacterial activity, *Candida spp*, concentration, efficacy, ethanol, flavonoid, Leafe, methanol, medicinal plants, microbial isolate, microorganisms, phytochemical, range, spectrum, tannin, discovery, technology, saponin, etc.

### Top cited articles with high Field Citation Ratio (FCR) and Relative Citation Ratio (RCR)

This study also revealed the top cited AMR articles indexed in Dimension database and sorted by FCR (Table 2). The FCR of the top 20 articles ranged from 4.09 to 36.54. The RCR ranged from 1.01 to 9.42. Table S2 shows the Nigeria’s top scholarly contributions to AMR indexed in Scopus database.

**Table 2:** Top 20 scholarly contributions to AMR authored by a Nigerian (Sorted by FCR)

| Authors | DOI | Year | Times cited | RCR | FCR |
| --- | --- | --- | --- | --- | --- |
| Stewardson et al | 10.1016/s1473-3099(18)30792-8 | 2019 | 92 | 8.45 | 36.54 |
| Olaitan et al | 10.1016/j.ijantimicag.2014.07.020 | 2014 | 203 | 9.42 | 24.66 |
| Nurjadi et al | 10.1093/jac/dku174 | 2014 | 75 | 3.13 | 19.42 |
| Shittu et al | 10.1186/1471-2180-11-92 | 2011 | 123 | 4.36 | 17.65 |
| Ghebremedhin et al | 10.1128/jcm.00648-09 | 2009 | 119 | 4.11 | 17.11 |
| Kesah et al | 10.1046/j.1469-0691.2003.00531.x | 2003 | 96 | 2.22 | 16.92 |
| Adesokan et al | 10.4102/ojvr.v82i1.816 | 2015 | 61 | 3.37 | 14.57 |
| Fashae et al | 10.3855/jidc.909 | 2010 | 79 | 2.56 | 11.58 |
| Aibinu et al | 10.1111/j.1469-0691.2011.03730.x | 2011 | 58 | 2.49 | 11.44 |
| Marin et al | 10.1371/journal.pntd.0002049 | 2013 | 82 | 3.23 | 9.96 |
| Titilawo et al | 10.1016/j.scitotenv.2015.03.095 | 2015 | 61 | 2.67 | 8.77 |
| Ogbolu et al | 10.1089/jmf.2006.1209 | 2007 | 77 | 1.01 | 8.09 |
| Ogbolu et al | 10.1016/j.ijantimicag.2010.08.019 | 2010 | 70 | 2.76 | 6.92 |
| Titilawo et al | 10.1007/s11356-014-3887-3 | 2015 | 63 | 2.73 | 6.21 |
| Akinyemi et al | 10.1016/j.puhe.2004.04.009 | 2005 | 53 | 1.15 | 5.98 |
| Okeke et al | 10.3201/eid0604.009913 | 2000 | 148 | 1.47 | 5.85 |
| Fortini et al | 10.1093/jac/dkr085 | 2011 | 71 | 2.76 | 5.69 |
| Chigor et al | 10.3390/ijerph7103831 | 2010 | 53 | 0.97 | 5.32 |
| Iwalokun et al | 10.1089/jmf.2004.7.327 | 2004 | 83 | 1.41 | 5.2 |
| Soge et al | 10.1093/jac/dki429 | 2005 | 64 | 1.83 | 4.09 |

## Discussion

AMR is an unfortunate result of antibiotic misuse and this study confirms that Institutions in Nigerian have contributed at least 0.2% of the global research output on AMR. A marked rise in scientific output started in 2016 and peaked in 2021. Although data for 2022 is incomplete, evidence of a continuing rise in research output is apparent. In general, the number of AMR documents in the database has gradually increased, implying a significant scientific research output. A similar upward trend has also been reported in earlier bibliometric studies[24–26]. This parallels the increased burden of AMR in Nigeria. Further, the marked rise in scientific output coincides with Nigeria’s response to a national rise in the burden of antimicrobial resistance that led to the development of the NAP (2017–2022) that sought to address crucial risk factors in harmony with the WHO Global Action Plan on AMR. Connecting the gap between research institutions, pharmaceutical industries and legislators is pertinent because the role of research institutions does not end with the production of scientific data. Research should translate to development of actionable and sustainable policies and practices. These findings also highlight the importance of strengthening research capacity across Nigerian institutions.

The majority of the documents (57.18%) were in the field of Medical and Health Sciences.

This result is comparable to (25,26), where the majority of research output was in a related field. There is a need for more research output in all subject areas, as well as more multidisciplinary research, to advance the science of AMR.

The centrality of an institution/organisation reflects its recognition in the field of AMR research[27]. By analysing both the centrality and number of articles, Nigeria’s AMR research strength was observed to be primarily focused at universities, research institutions and NGOs. The University of Ibadan was Nigeria’s leading research institution in terms of scientific outputs and centrality. International Institute of Tropical Agriculture was the leading nongovernmental organization (NGO) in terms of centrality and Nigerian Institute of Medical Research lead in scholarly output among research institutes.

We found that OKEKE IN ranked first in publication frequency and second to BAUER AW (0.13) in terms of centrality (0.07), indicating that the author was part of an influential core author group in the field of AMR research in Nigeria. Majority of Nigerian authors had a centrality index of less than one. This result indicated that more researchers should participate actively in studies on AMR burden and contribute important scientific input to this field.

African Journal of Clinical and Experimental Microbiology was ranked as the top journal, publishing AMR documents with a minimum of one Nigerian institutional affiliation. The same journal was also the most cited in terms of published documents. Although Nigerian journals were among the top publishers of AMR research featuring at least a Nigerian author, there is a need to invest in strengthening local journals in Nigeria in order to achieve comparable indexing status and scientific standards to those of the world’s top journals.

The frontier in a research area depicts the current growth path of a discipline in bibliometrics, and thus the references in the frontier article represent the field’s intellectual core[28]. We described Nigeria’s scholarly core in the field of AMR by mapping references. Themes on antimicrobial stewardship, Clinical and laboratory practice, public health implication, traditional/molecular methods, and phytomedicine and drug discovery are major research trends of Nigeria’s scientific community. These thematic distributions align with the observations of [29]. It is pertinent to boost research in all thematic areas of AMR research in the country to significantly facilitate processes that will slow down the spread of AMR in the country. Research themes exploring the use of molecular and AI systems for AMR detection, surveillance, and drug discovery must be enhanced to confront rising bacterial resistance by significantly reducing the time taken to find new antimicrobials, improving diagnostic and treatment precision, as well as lowering costs[30]. A hotspot is a scientific topic of discussion in a series of documents which are inextricably connected to a particular time period[31]. The keywords are generalizations of the literature’s topic[31]. A high-frequency keyword analysis can be used to identify hotspots in the field of AMR research. From our results “*Escherichia coli*” and “*Staphylococcus aureus*”, “ESBL”, “*Salmonella*”, “tetracycline” and “Plasmid” were the most frequently used keywords. Nigerian researchers could conduct advanced studies based on the frequently discussed topics in this field.

An FCR range of 4.09 to 36.54 reflect the moderate to high scientific influence of Nigerian researchers in AMR research field. The theme of the top-cited documents in both the dimension and Scopus databases discussed are similar. The articles were focused on key AMR issues and determinants of human and animal importance relevant to the Nigerian and African population. Including; infectious diseases, Clinical and community health. Specifically, they highlighted the Prevalence and/or Emergence of colistin resistance *Klebsiella pneumoniae*, Community-Associated Methicillin-Resistant *Staphylococcus aureus*, carbapenem resistance, Enterobacteriaceae, Multidrug Resistant Non-O1/Non-O139 *Vibrio cholerae*, trimethoprim resistance gene *dfr*G in *Staphylococcus aureus*, Plasmid-mediated quinolone resistance and betalactamases in *Escherichia coli*, CTX-M-15 extended-spectrum Beta-lactamase from *Klebsiella pneumoniae* etc. The identification of top-cited articles is useful as it reflects the diverse scope and health research fields that are of high interest to the readership of AMR, in addition to helping researchers identify the authors and institutions that have highly contributed to such influential studies for future research collaborations.

We are unlikely to witness a return to the heyday of antibiotic discovery, when numerous chemotypes were being regularly discovered and brought into clinical use in a short time frame. What does the antibiotic future hold? An assortment of approaches to infection treatment and prevention will have to be offered in the future. Anti-virulence strategies, Small-molecule antibiotics, peptide-based antibiotics, Bacteriophage Monoclonal antibodies, Probiotic strategies and Vaccines are future trends in AMR and drug discovery.

Increasingly, the traditional Waksman platform returns recognized molecules. with recent breakthroughs in analytical chemistry, synthetic biology, genome mining, and genomics, the spectacular chemical diversity of nature can be taken advantage of by moving beyond the traditional Waksman platform. For instance, many environmental bacteria like actinomycetes harbour genetic programs to produce 20 to 40 natural products, while fungi can contain even more[32]. Avoiding actinomycetes bacteria is one strategy, for example. Some studies have identified 2 new antibiotics with new mechanisms of action, teixobactin, from the betaproteobacterium *Eleftheria terrae* isolated from a soil specimen[33], and darobactin, from the gamma proteobacterium *Photorhabdus khaini* isolated from the gut of insect-killing nematodes[34].

The Myers laboratory has developed innovative modular pathways for the synthesis of new therapeutic tetracycline, macrolide, and lincosamide antibiotics, significantly expanding the diversity of these compound classes well beyond limits of natural biosynthesis[32]. Artificial intelligence approaches could also mine chemical libraries for therapeutic potential in order to develop new synthetic scaffolds[30].

Despite the difficulties in bringing peptide-based antibiotics to the clinic (most suffer from haemolytic activity or nephrotoxicity), optimism remains, and many AMPs are in various stages of development, targeted toward a range of pathogens[32].

Though naturally narrow in spectrum and must be combined with antibiotics, Antivirulence strategies aimed at abrogating pathogen virulence are a future trend. Sibofimloc, which prevents pathogenic *E. coli* from adhering to host epithelia is entering phase 2 trials[35].

Adjuvants or antibiotic combinations, such as b-lactamase inhibitors, can aid in the fight against resistance. A combo of bedaquiline, pretomanid, and linezolid, for instance, was recently approved to treat of Extensively drug - resistant M. tuberculosis, and more combinations are being studied[36].

In current history, the effectiveness and narrow spectrum of phage have been acknowledged as a valuable resource in the treatment of highly drug-resistant infections. The concept of using bacterial phages to treat drug-resistant infections is becoming more appealing. Phage therapy is increasingly being viewed as an alternative to conventional antibiotics in medicine, spurring more clinical trials, the majority of which are in phases one and two [32].

There are currently only 3 approved antimicrobial Monoclonal antibodies (mAbs), two of which inhibit the virulence of Bacillus anthracis (raxibacumab and obiltoxaximab), and one of which inhibits the virulence of *Clostridium difficile* (bezlotoxumab), reducing the incidence of reoccurrence. Only bezlotoxumab withstood clinical trials in its entirety[37], whereas the raxibacumab and obiltoxaximab were approved under the animal rule for biothreat organisms. But besides their proven effectiveness, few are now in late-stage clinical trials (phase 2, *P. aeruginosa*: panobacumab/Aerumab; phase 2, *S. aureus*: omodenbamab/514G3 and; phase 3, *S. aureus*: suvratoxumab and tosatoxumab[32].

### Limitations

The data of this study were mainly derived from the dimensions database on August 20^th^ 2022, and other literature databases like Web of Science, Open Alex and Semantic scholar were not searched. Therefore, we might not have been able to retrieve all papers on this topic, and selection bias may have been present.

## Conclusion and Recommendations

According to this research, Nigerian institutions and organizations contribute moderately to AMR research output. However, there is a pressing need to strengthen the country’s research capacity and expedite the use of scientific insights provided by its scientists across all subject areas in order to make a significant contribution to development.

Nigeria is home to more than 206,139,589 people (an annual change of 2.58%)[38,39], and effective research is crucial to the advancement of the country. According to our research, Nigeria’s contribution to the world’s AMR scientific output was only 0.2%. This is scanty, and it emphasizes that AMR response is not always tied to local data, instead it relies on adapting global policies and those of global health organizations. Developing local data to better understand AMR trends is still an excellent method for Nigeria to respond to health emergencies. Nigeria’s potential to respond to health issues needs to be strengthened, and investing in research is one excellent way to do so.

Federal and state academic universities, which form the bulk of academic research institutions in Nigeria, have been on strike for over 65 months since the country’s independence in 1999. This illustrates that Nigeria’s academic institutions are underrated as significant contributors to the nation’s information economy and growth plans. A university ought to be a valuable source of knowledge that helps with innovation and national growth[39]. The combined effect of resistance pressures and a lack of creativity and determination to develop next-generation therapies threatens modern medicine’s very existence. Artificial intelligence (AI) frameworks are not being utilized in the country. Whole-genome sequencing for antimicrobial susceptibility testing (WGS-AST) is used to diagnose AMR with high accuracy and consistency by extracting large and high-dimensional datasets. AI systems are used to improve this process. Fundamentally, the antimicrobial resistance (AMR) problem is one of kinetics: While the development of antimicrobials is slow, resistance develops quickly. AI technologies have the potential to ease synthesis of antimicrobial peptides with machine/deep learning, New and cost-effective antibiotics, appropriate antibiotic prescription, prediction of antibiotic resistance, prediction of the severity of infection (efficiency of distinguishing infectious and non-infectious diseases) and by extension, Mortality reduction. Nigeria should adopt current AI practices regarding antibiotic resistance across the world, with Nigeria as the central focus.

The burden of AMR is Monumental and existential and the Right priority ought to be given towards achieving Sustainable standards in the fight against it. Evidence-driven economy is imperative and national stakeholders must be devoted to backing education by adapting the United Nations Educational, Scientific and Cultural Organizations (UNESCO) recommendations[40]. Investment in improving the student’s professor ratio in Nigerian Universities will aid in improving the quality of scientific output. New antibiotics are always welcomed, the big question is, where to find these candidates? Nigerian universities must be revitalized so that they can meet the country’s development needs and herald a generation of patriots with impactful knowledge.

## Data Availability

All data produced in the present study are available upon reasonable request to the authors

